# Diffuse fibrosis, coronary microvascular dysfunction and systolic dysfunction in Wilson disease

**DOI:** 10.1101/2024.10.11.24315326

**Authors:** Rebecka Steffen Johansson, Csenge Fogarasi, Peter Kellman, Andreas Kindmark, Jannike Nickander

## Abstract

**Background:** Wilson disease (WD) causes intracellular copper accumulation in the body due to a genetic defect in the protein ATP7B. Cardiac involvement such as electrocardiographic abnormalities, rhythm abnormalities, heart failure and cardiac death have been reported, however pathophysiological mechanisms remain unclear.

**Objectives:** This study aimed to comprehensively assess the myocardium in WD patients without cardiac symptoms using multiparametric cardiovascular magnetic resonance imaging (CMR), including quantitative stress perfusion mapping and strain analysis.

**Methods:** WD patients (n=17, 41±16 years, 47% female) and volunteers (n=17, 39±15 years, 47% female) underwent multiparametric mapping at 1.5 T CMR including cine, native T1, native T2, adenosine stress perfusion mapping, late gadolinium enhancement (LGE), and extracellular volume (ECV) imaging. Symptoms of myocardial ischemia were quantified using Seattle Angina Questionnaire-7 (SAQ-7) and cardiovascular risk factors and medications were recorded.

**Results:** Both stress perfusion and MPR were lower in WD patients (2.95±0.58 vs 3.67±1.01 ml/min/g, and 3.4±0.8 vs 4.4±1.9), while ECV was higher, (29±3% vs 27±2%), p<0.05 for all. Left ventricular ejection fraction (LVEF) was lower in WD patients, (56±6% vs 61±6%, *p*=0.02), and LV ventricular global circumferential strain (LV GCS) was higher (-18±2% vs - 20±2%, *p*=0.005). Late gadolinium enhancement (LGE) was present in the right ventricular insertion point (RVIP) in 12/17 (71%) of the WD patients.

**Conclusions:** In this small mechanistic study, WD patients on stable treatment without apparent cardiac symptoms have early signs of diffuse fibrosis, coronary microvascular dysfunction (CMD) and systolic dysfunction, shedding light on pathophysiological mechanisms of cardiac dysfunction in copper accumulation.

## Introduction

Wilson disease (WD) is a rare autosomal recessive genetic disease affecting the copper-transporting protein ATP7B, causing copper accumulation primarily in the liver and brain (1). Cardiac involvement in WD was described already during the 1980s, however has received relatively little attention (2). WD patients have higher risk of atrial fibrillation (AF) and congestive heart failure (HF), adjusted for age, sex and cardiovascular risk factors and presents with HF earlier than non-WD patients (3). Even in young WD patients, electrocardiographic (ECG) abnormalities, rhythm disturbances, cardiac hypertrophy and subclinical systolic and diastolic dysfunction have been shown (4–6). Although rare, cardiac death in young WD patients have occurred (7). Therefore, it has been suggested that cardiac involvement should be evaluated in WD (8). While insufficiently studied in the myocardium, copper may cause oxidative stress, leading to necrosis, inflammation and fibrosis in the liver (7). Therefore, comprehensive multiparametric cardiovascular magnetic resonance imaging (CMR) enabling tissue characterization may provide insights into the pathophysiology and phenotype of cardiac WD. Moreover, CMR may simultaneously assess cardiac anatomy and function, quantify perfusion and provides prognostic information (9). There are no prior studies comprehensively assessing cardiac involvement in WD patients without cardiac symptoms, using multiparametric CMR including quantitative adenosine stress perfusion mapping. Therefore, the current study aimed to assess the myocardium in WD patients without cardiac symptoms, compared to age- and sex-matched healthy volunteers, using comprehensive multiparametric CMR including quantitative adenosine stress perfusion mapping and strain analysis.

## Methods

### Study group

Patients with established WD diagnosis (Leipzig score ≥4) (10), clinically stable with regards to symptoms and laboratory workups (blood counts, liver and kidney function) on WD medication for at least 1 year, were identified and invited to participate in the study at the endocrinology clinic at Uppsala University Hospital, Sweden. Liver affliction dominated in 11 patients, 5 had neurological WD. Most were treated with the chelator trientine (n=9), or zinc (n=4). One WD patient had a liver transplant and two were untreated due to mild disease, Table 1. Exclusion criteria were standard contraindications to CMR and adenosine such as severe asthma, decreased renal function (serum creatinine >150 μmol/l), pacemaker, or claustrophobia. WD patients were examined between January 2022 and March 2023. Sex- and age-matched healthy volunteers without cardiovascular symptoms who underwent CMR between 2016 and 2022 were included for comparison (11, 12). Figure 1 shows the flow-chart of the inclusion process. An ECG and history of cardiovascular risk factors and medications were obtained prior to CMR. The Seattle Angina Questionnaire 7 (SAQ-7) was used to quantify cardiac symptoms and disability, and results summarized as the SAQ-7 summary score (13). The Swedish Ethical Review Authority granted ethical approval for the study (DNR 2019-06287, with amendments 2019-06287, 2021-05508-02, 2022-06995-02 and for volunteers 2015/2106–31/1 and 2014/131-31/1, 2017/2415-32/1, 2021-06837-02). All participants provided written informed consent.

**Figure 1.**
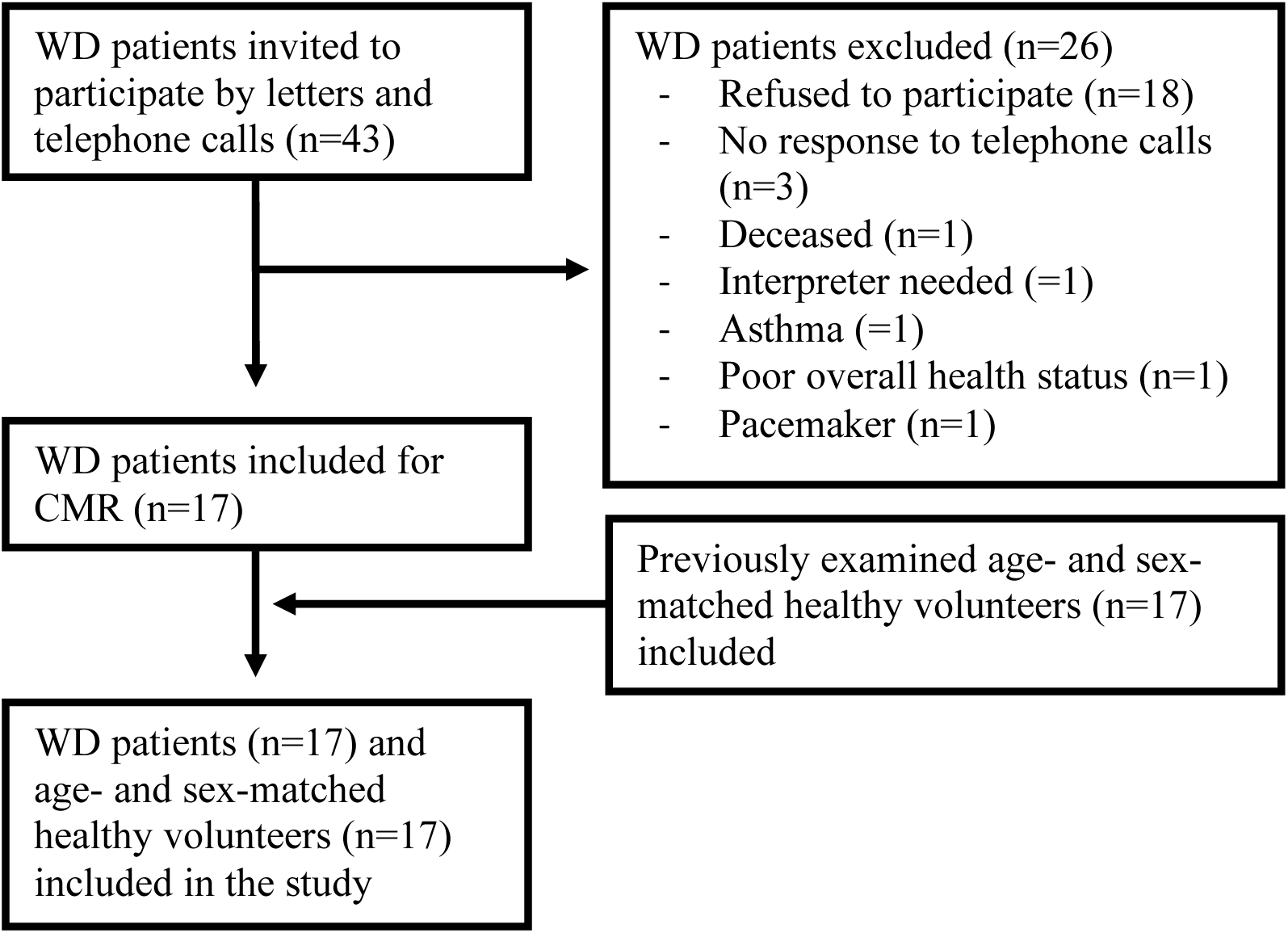
Flow chart of study cohort. The figure shows inclusion and exclusion into the study. Abbreviations: CMR = cardiovascular magnetic resonance imaging; WD = Wilson disease.

**Table 1.** Clinical characteristics of WD patients.

|  | **All (n=17)** | **Male (n=9)** | **Female (n=8)** |
| --- | --- | --- | --- |
| **Age at CMR (years)** | 41±16 | 36±14 | 46±17 |
| **Disease duration at CMR (years)** | 24±14 | 18±13 | 30±13 |
| **<u>Main WD manifestation, n</u>** |  |  |  |
| **None** | 1 | 0 | 1 |
| **Neurological** | 5 | 2 | 3 |
| **Liver** | 11 | 7 | 4 |
| **<u>Treatment, n</u>** |  |  |  |
| **None** | 2 | 0 | 2 |
| **Zinc** | 4 | 1 | 3 |
| **Penicillamine** | 1 | 1 | 0 |
| **Trientine** | 9 | 6 | 3 |
| **Liver transplant** | 1 | 1 | 0 |

Age at CMR and disease duration at CMR presented as mean ± standard deviation in years, as well as main WD manifestation and treatment presented as n, for all and divided by sex. Abbreviations: WD = Wilson disease; CMR = cardiovascular magnetic resonance.

### Image acquisition

CMR images were acquired supine with a Magnetom Aera® 1.5 Tesla (T) scanner (Siemens Healthineers, Erlangen, Germany), together with a phased-array 18-channel body matrix coil and a spine matrix coil. Venous blood was drawn prior to imaging to determine hematocrit and creatinine. Cine images in standard three long-axis and short-axis slices were obtained by full coverage retrospective ECG-gated balanced steady state free precession (bSSFP) cine imaging.

Quantitative perfusion maps (ml/min/g) were obtained by first pass perfusion imaging in three short-axis slices (basal, midventricular, apical) during administration of intravenous contrast (bolus 0.05 mmol/kg, gadobutrol, Gadovist, Bayer AB, Solna, Sweden). Perfusion maps were acquired in adenosine stress (infusion 140 µg/kg/min or if insufficient adenosine response according to symptoms or heart rate response increased according to clinical routine to 210 µg/kg/min (Adenosin, Life Medical AB, Stockholm)) and in rest. The subjects abstained from caffeine for 24 hours and nicotine 12 hours before the CMR. Two different cannulas were used for administration of gadobutrol and adenosine. Perfusion maps were computed using a distributed tissue exchange model (14) and generated using the Gadgetron inline perfusion mapping software (15).

Native T1 maps were acquired in three short-axis slices with an ECG-gated Modified Look-Locker Inversion Recovery (MOLLI) prototype sequence. Post-contrast T1 maps were acquired with the same slice position, after administration of intravenous contrast (bolus in total 0.2 mmol/kg, gadobutrol). Extracellular volume (ECV) maps were generated from the native and post-contrast T1 maps and calibrated by the hematocrit (16, 17). Native T2 maps were obtained with a T2-prepared sequence (Siemens MyoMaps product sequence) in three short-axis slices. Late gadolinium enhancement (LGE) images were obtained in standard three long-axis and short-axis slices following contrast administration, with a free breathing phase-sensitive inversion recovery (PSIR) sequence with bSSFP single shot readout.

### Image analysis

Images were anonymized and analyzed offline with the CMR analysis software Segment (vers 3.3 R9405e Medviso AB, Lund, Sweden) (18). Left ventricle (LV) mass (LVM) and volumes were obtained by delineating endo- and epicardial contours in end-systole and end-diastole in the short-axis cine stack. Right ventricle (RV) volumes were obtained by delineating RV endocardial contours in end-systole and end-diastole in the short-axis cine stack. Left ventricular global longitudinal strain (LV GLS), global circumferential strain (LV GCS) and global radial strain (LV GRS) were acquired using the feature-tracking strain module following delineation of endocardial contours in the cine long-axis slices and endo- and epicardial contours in the short-axis stacks in end-diastole (19, 20). Right ventricular global longitudinal strain (RV GLS) was likewise acquired using the feature-tracking strain module following delineation of the RV endocardial contours in end-diastole in the cine four chamber long-axis slice. RV and LV atrioventricular plane displacement (RV AVPD, LV AVPD) were assessed using the relevant module in Segment.

Native T1, native T2, ECV and perfusion maps were analyzed by delineating the endo- and epicardial contours of the LV in each short-axis stack and values exported in a 16-segment model (21). A 10% erosion margin was used for endo- and epicardial contours to avoid partial volumes effects. LGE images were evaluated with regards to focal fibrosis. Liver T1 values were obtained by placing three regions of interest over the liver in the midventricular native T1 map, avoiding vessels and surrounding tissues, and averaging the values.

Native T1, native T2, ECV, perfusion maps, and cine images were assessed for inter- and intra-observer variability, the former in all 17 WD patients by two independent observers and the latter in 10 patients by one observer.

### Statistical analysis

Continuous data were presented as mean ± standard deviation (SD) or median [interquartile range] and categorical data as numbers (percentages). Normality was assessed using the Shapiro-Wilk test. Mass and volumes were indexed to body surface area (BSA) calculated by Mosteller formula (22). Global native T1, native T2, ECV and perfusion values were obtained by averaging segmental values. Segments with LGE were excluded from the ECV analysis. Rate pressure product (RPP) was calculated as heart rate multiplied by systolic blood pressure. Myocardial perfusion reserve (MPR) was calculated as stress perfusion divided by rest perfusion. Continuous data were compared using the independent t-test or Mann-Whitney U test as appropriate. Categorical data were compared using Fisher’s exact test. The relationships between native T1, ECV, native T2, LV GCS, LV GRS and LV GLS with stress perfusion and MPR were explored using multiple linear regression. Inter- and intra-observer agreement was calculated for global native T1, native T2, ECV, rest and stress perfusion, MPR, LVEF and LV GCS, LV GRS and LV GLS. The intra-class correlation coefficient (ICC) was good or excellent (0.82-1.00, *p*<0.05 for all, data shown in Table 1 in Appendix). Previously published data on healthy volunteers show 16 participants needed to detect a 0.78 ml/min/g difference in stress perfusion with power 80% and alpha 0.05 (23). Data was gathered in Microsoft Excel (Microsoft, Redmond, Washington, USA) and statistical analysis performed in Excel and IBM SPSS Statistics (IBM SPSS Statistics version 28, IBM, New York, USA). The significance level was defined as *p*<0.05.

## Results

### Clinical characteristics

Both groups comprised of 17 individuals, of which 8 (47%) were female, WD patients were aged 41±16 years and volunteers 39±15 years. Except for slightly higher hematocrit in the volunteers, there were no differences in clinical characteristics including body composition, cardiovascular risk factors and medications, Table 2. Additionally, no one had a history of hypertension or treatment with angiotensin-converting-enzyme inhibitors or angiotensin receptor blockers, hyperlipidemia or statin treatment, or were current smokers. One WD patient had angina pectoris, treated with diltiazem. ECG showed sinus tachycardia (heart frequency 101 beats per minute) in one WD patient, a small intraventricular conduction defect in one patient and sinus rhythm with frequent supraventricular and ventricular extra beats in one patient. Otherwise, ECGs were normal (ECG missing in 3 patients). SAQ-7 summary score for the WD patients was 100 [95 - 100] (data missing for 2 patients).

**Table 2.** Clinical characteristics at CMR of WD patients and volunteers.

| Clinical characteristics | WD patients (n=17) | Volunteers (n=17) | *p* |
| --- | --- | --- | --- |
| Female sex, n (%) | 8 (47%) | 8 (47%) | 1.00 |
| Age (years) | 41±16 | 39±15 | 0.80 |
| Height (cm) | 176±10 | 175±11 | 0.79 |
| Weight (kg) | 77±14 | 70±14 | 0.13 |
| BSA(m<sup>2</sup>) | 1.9±0.2 | 1.9±0.2 | 0.19† |
| Creatinine (mmol/l) | 77±16 | 70±14 | 0.12† |
| Hematocrit (%) | 40±3 | 43±3 | **0.02** |
| Angina pectoris, n (%) | 1 (6%) | 0 (0%) | 0.49 |
| Diabetes mellitus, n (%) | 1 (6%) | 0 (0%) | 0.49 |
| Smoking previously, n (%) | 5 (31%) | 7 (41%) | 0.72 |
| Beta blockers, n (%) | 1 (6%) | 0 (0%) | 0.49 |
| Calcium channel blockers, n (%) | 1 (6%) | 0 (0%) | 0.49 |

Clinical characteristics are presented as n (%) where p-values denotes the Fisher's exact test or as mean ± standard deviation where p-values denotes the independent t-test and p-values marked † denotes the Mann Whitney U test. Abbreviations: BSA = body surface area; WD = Wilson disease.

### CMR findings

#### Multiparametric CMR mapping

Multiparametric CMR mapping results are shown in Table 3. There were no differences in global native myocardial or hepatic T1, or in global native myocardial T2. Global ECV was higher in the WD patients compared to volunteers (29±3% vs 27±2%, *p*=0.003). Examples of native T1, T2 and ECV maps of a WD patient and a healthy volunteer are shown in Figure 2.

**Figure 2.**
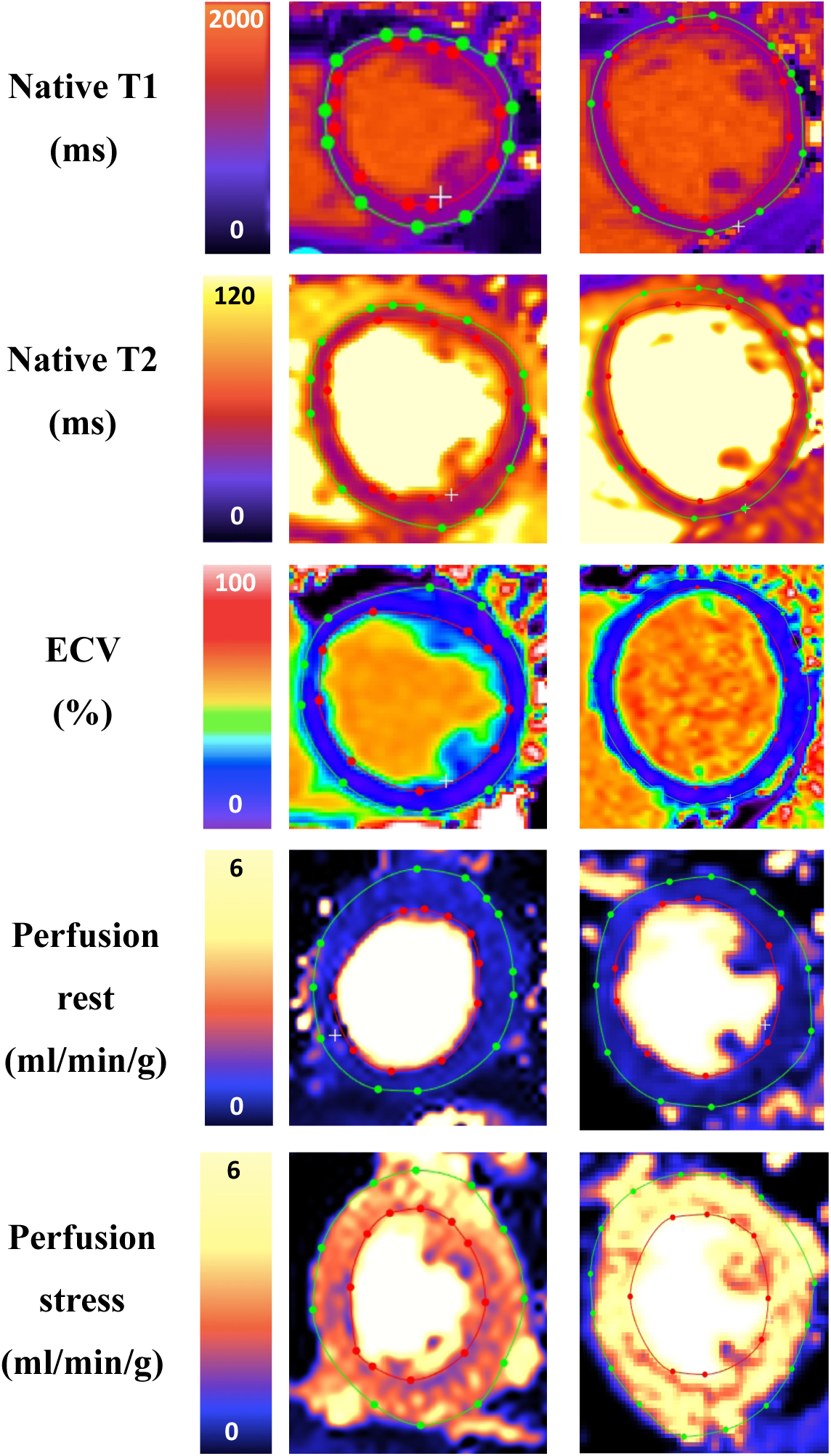
Multiparametric CMR in a WD patient and volunteer. Examples of midventricular short-axis native T1 (ms), native T2 (ms), and ECV (%), rest and stress perfusion maps (ml/min/g) maps, including segmentation delineating endocardial (red) and epicardial (green) borders. Abbreviations: ECV = extracellular volume, WD = Wilson disease.

**Table 3.** Multiparametric CMR mapping.

| **CMR findings** | **WD patients (n=17)** | **Volunteers (n=17)** | ***p*** |
| --- | --- | --- | --- |
| **Native T1 myocardium (ms)** | 995±41 | 992±36 | 0.67 |
| **Native T1 liver (ms)** | 571±101 | 556±45 | 0.46† |
| **Native T2 (ms)** | 49±3 | 49±2 | 0.49 |
| **ECV (%)** | 29±3 | 27±2 | **0.003** |

CMR findings are presented as mean ± standard deviation. P-values denotes the independent t-test, p-values marked † denotes the Mann Whitney U test. Abbreviations: WD = Wilson disease; CMR = cardiovascular magnetic resonance imaging; ECV = extracellular volume.

#### Quantitative stress perfusion mapping

Perfusion maps are missing in one WD patient due to contrast failure. Stress perfusion and MPR were lower in WD patients compared to volunteers (2.95±0.58 vs 3.67±1.01 ml/min/g, *p*=0.017 and 3.4±0.8 vs 4.4±1.9, *p*=0.044), but there was no difference in rest perfusion, Table 4 and Figure 3. Examples of midventricular perfusion maps in stress and rest of a WD patient and a healthy volunteer are shown in Figure 2. Using multiple linear regression, native T1, ECV, native T2, LV GCS, LV GRS and LV GLS combined did not correlate with stress perfusion or MPR in WD patients, nor did native T1, ECV and native T2 combined correlate with stress perfusion or MPR in the patients (data not shown, model *p*>0.05 for all). LV GCS, LV GRS and LV GLS combined did not correlate with stress perfusion or MPR in patients. However, LV GCS and LV GLS correlated with stress perfusion in WD patients (LV GCS β -0.62, *p*=0.033, and LV GLS β 0.67, *p*=0.024, model R^2^ 0.38, *p*=0.045), but not with MPR (data not shown, model *p*>0.05).

**Figure 3.**
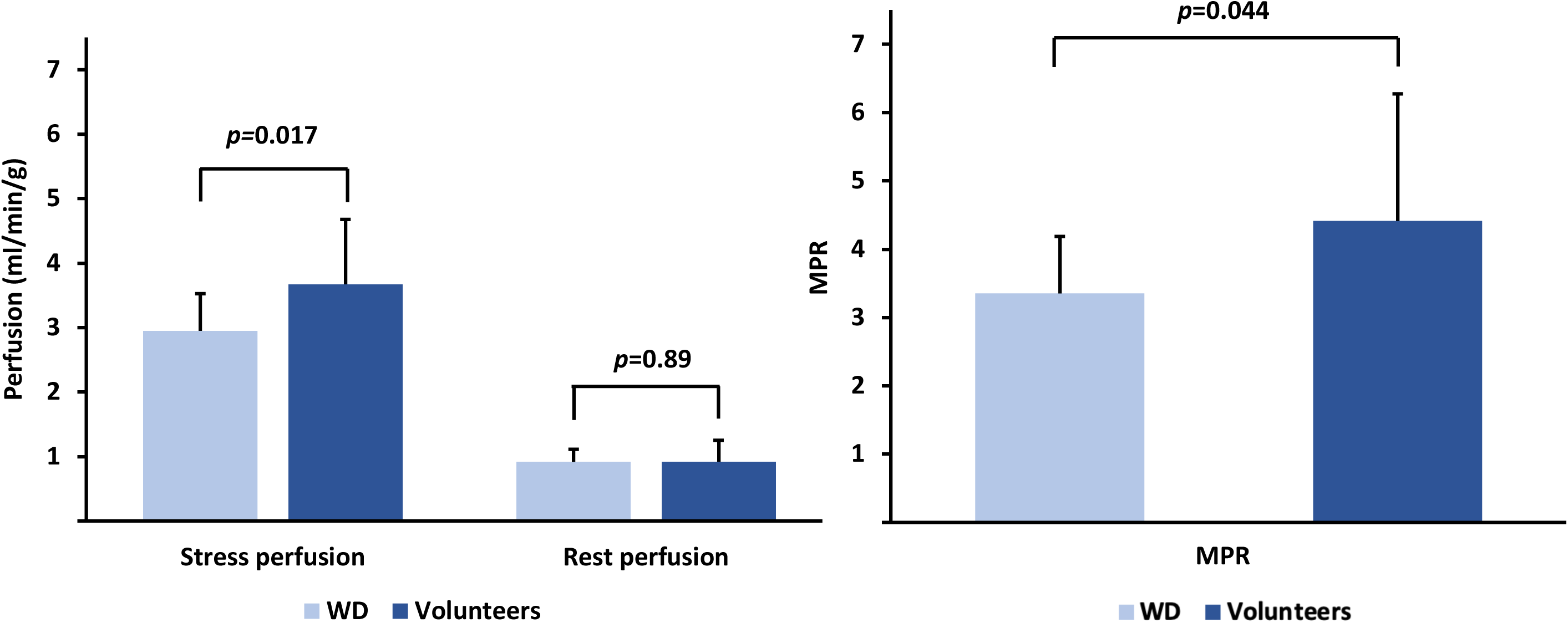
Stress and rest perfusion and MPR in WD patients and volunteers. Bar charts show mean, and error bars show standard deviation of stress and rest perfusion (ml/min/g) and MPR in WD patients and volunteers, together with p-values. Abbreviations: WD = Wilson disease; MPR = myocardial perfusion reserve.

**Table 4.** Stress perfusion CMR mapping.

| CMR findings | WD patients (n=16) | Volunteers (n=17) | *p* |
| --- | --- | --- | --- |
| Heart rate rest (bpm) | 73±11 | 70±9 | 0.32 |
| SBP rest (mmHg) | 122±15 | 111±9 | **0.032**† |
| RPP rest | 8895±1912 | 7583 ± 814 | 0.06† |
| Heart rate stress (bpm) | 93±18 | 93 ± 12 | 0.89 |
| SBP stress (mmHg) | 119±12 | 115±14 | 0.49 |
| RPP stress | 11131±2681 | 10733±2158 | 0.64 |
| Perfusion stress (ml/min/g) | 2.95±0.58 | 3.67±1.01 | **0.017** |
| Perfusion rest (ml/min/g) | 0.92±0.20 | 0.92±0.33 | 0.89† |
| MPR | 3.4±0.8 | 4.4±1.9 | **0.044** |

CMR findings are presented as mean ± standard deviation, p-values denotes the independent t-test, p-values marked † denotes the Mann Whitney U test. Perfusion maps in rest and stress are missing in one WD patient due to contrast failure, and heart rate, blood pressure and RPP in rest and stress were therefore only assessed in 16 WD patients. Abbreviations: WD = Wilson disease; CMR = cardiovascular magnetic resonance imaging; SBP = systolic blood pressure; RPP = rate pressure product; MPR = myocardial perfusion reserve.

#### LGE

LGE was present in the right ventricular insertion point (RVIP-LGE) in 12/17 (71%) of WD patients (Figure 4A). One WD patient had basal inferolateral non-ischemic midmural LGE (Figure 4B), with native T2 52 ms in the scar. One WD patient had basal inferolateral non-ischemic midmural and epicardial LGE (Figure 4C), with native T2 59 ms in the scar. One WD patient had basal lateral non-ischemic midmural LGE (Figure 4D), with native T2 57 ms in the scar, and this individual had frequent supraventricular and ventricular extra beats. Two WD patients had no LGE.

**Figure 4.**
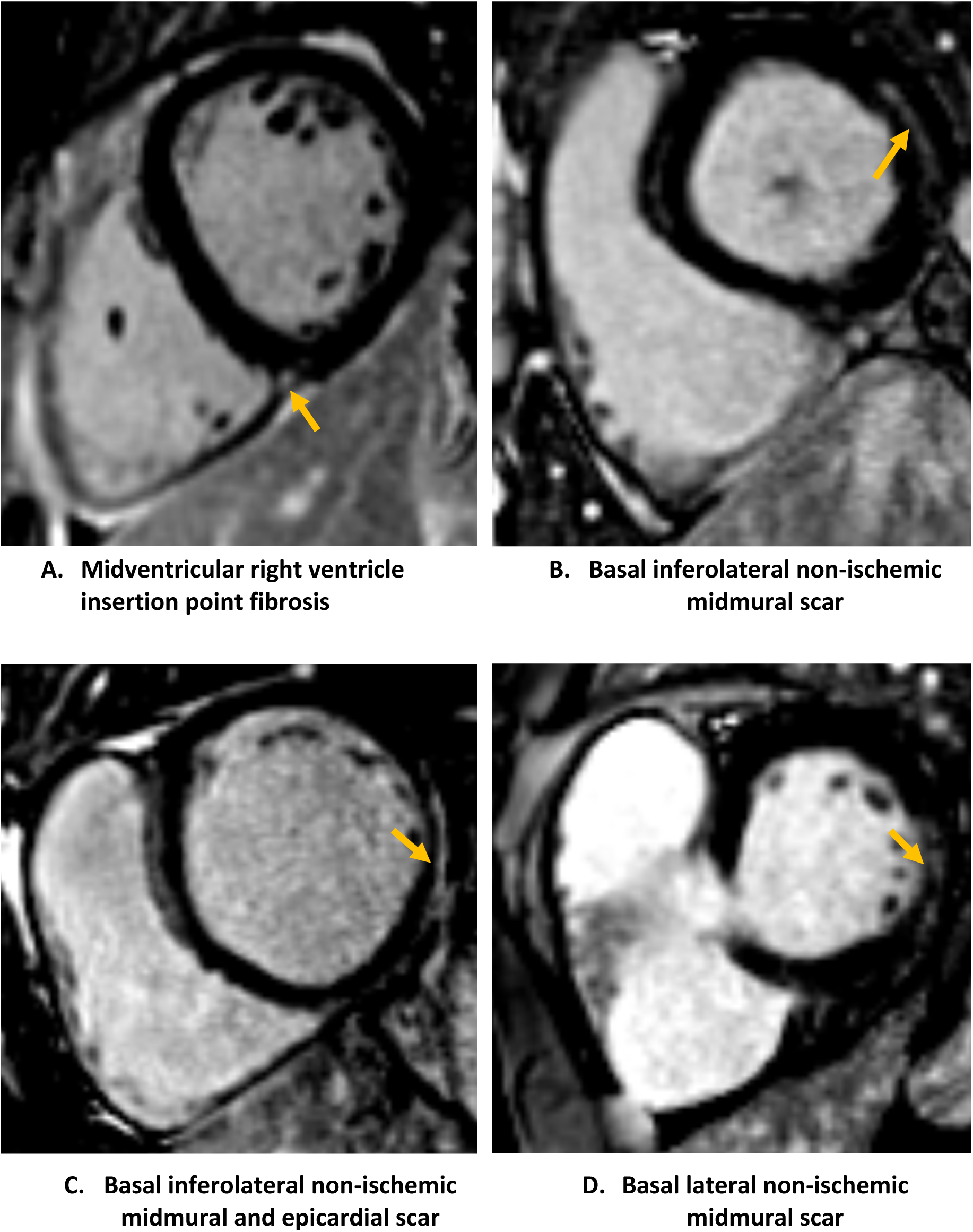
LGE examples in WD patients, indicated by arrows. Figure 4A shows midventricular RVIP-LGE. Figure 4B-4D show non-ischemic scars. Abbreviations: LGE = late gadolinium enhancement; WD = Wilson disease, RVIP = right ventricular insertion point.

#### Cardiac function

WD patients had lower left ventricular ejection fraction (LVEF) compared to volunteers (56±6% vs 61±6%, *p*=0.02), but no other differences were found in LV mass or volumes, Table 5. LV GCS was higher in WD patients compared to volunteers (-18±2% vs -20±2%, *p*=0.005), but no other differences were found in LV GRS, LV GLS, or LV AVPD. There were no differences in RV volumes, RV GLS and RV AVPD (data shown in Table 2 in Appendix).

**Table 5.** LV volumes and function.

| CMR findings | WD patients (n=17) | Volunteers (n=17) | *p* |
| --- | --- | --- | --- |
| LVM (ml) | 99±28 | 95±22 | 0.57 |
| LVM (g) | 95±30 | 100±23 | 0.57 |
| LVMi (g/m<sup>2</sup>) | 48±12 | 54±9 | 0.15 |
| LVEDV (ml) | 190±60 | 176±46 | 0.45 |
| LVEDVi (ml/m<sup>2</sup>) | 98±25 | 94±17 | 0.66 |
| LVESV (ml) | 85±38 | 68±22 | 0.22† |
| LVESVi (ml/m<sup>2</sup>) | 43±16 | 37±9 | 0.29† |
| LVSV (ml) | 105±28 | 108±28 | 0.77 |
| LVSVi (ml/m<sup>2</sup>) | 54±12 | 58±11 | 0.39 |
| LVEF (%) | 56±6 | 61±6 | **0.02** |
| CO (l/min) | 7.5±3 | 7.1±1.8 | 0.93† |
| LV GLS (%) | -18±2 | -19±2 | 0.22† |
| LV GCS (%) | -18±2 | -20±2 | **0.005†** |
| LV GRS (%) | 40±2 | 44±2 | 0.23 |
| LV AVPD (mm) | -14±2 | -15±2 | 0.34 |

CMR findings are presented as mean ± standard deviation. P-values denotes the independent t-test, p-values marked † denotes the Mann Whitney U test. Abbreviations: WD = Wilson disease; LV = left ventricle; CMR = cardiovascular magnetic resonance imaging; LVM = Left ventricular mass; LVEDV = left ventricular end-diastolic volume; LVESV = left ventricular end-systolic volume; LVSV = left ventricular stroke volume; LVEF = left ventricular ejection fraction; CO = Cardiac output; LV GLS = left ventricular global longitudinal strain; LV GCS = left ventricular global circumferential strain; LV GRS = left ventricular radial strain; LV AVPD = left ventricular atrioventricular plane displacement. Values marked i are indexed to body surface area (BSA) according to the Mosteller formula.

## Discussion

To the best of our knowledge, this is the first study investigating cardiac involvement in WD using comprehensive CMR, including quantitative stress perfusion mapping. While the current study is small, we showed RVIP-LGE, higher global ECV indicating early diffuse fibrosis, lower stress perfusion and MPR indicating CMD, and mildly impaired LV GCS and lower LVEF indicating early cardiac dysfunction, despite no cardiac symptoms.

### Multiparametric CMR mapping in WD

Previous CMR studies in WD are few and show conflicting results, likely due to differences in sex, age, ethnicity, sample size, and importantly – disease stage and duration of copper exposure, and results of decoppering. Copper is paramagnetic and does not affect signal intensity (SI); the SI changes seen in brain magnetic resonance imaging in WD are rather thought to be secondary to edema or necrosis (24). Elevated native T1 and T2 reflect active inflammation, edema or necrosis, caused by infiltration or systemic disease (25). Global native T1 and T2 were unaffected in the current study. In contrast, previous studies have shown increased native T1, native T2 and ECV with normal LVM, biventricular volumes and strain (26, 27), both without LGE (26) and with RVIP-LGE (27). Elevated native T1 and ECV with normal native T2 have also been shown in WD, with higher native T1 and ECV in neurological WD (28). LGE was also shown, particularly in the RVIP and interventricular septum, and HF was present in neurological WD (28). Similarly, ECV was elevated, and we also demonstrated RVIP-LGE. Moreover, three WD patients had non-ischemic LGE of which two had elevated native T2 in the scar, possibly indicating active focal inflammation despite no current exacerbation. Interestingly, elevated native T2 with myocarditis-pattern has been shown in WD, together with RV dysfunction and LGE, and in WD with acute exacerbation native T2 normalized following decoppering (29). We theorize that elevated native T1 and T2 may reflect more acute or severe WD, particularly as neurological WD is considered more advanced with longer disease duration (30). Subgroup analysis comparing neurological and non-neurological WD was not performed in the current study due to small sample size. Moreover, native liver T1 did not differ between WD patients and volunteers and was within normal range (31). In liver cirrhosis, it has previously been shown increased native T1, T2 and ECV in both liver and heart, with LGE and impaired LV GLS (32). Thus, this indicates hepatic and myocardial inflammation and fibrosis with functional impairment, not yet present to such an extent in the current study with stable WD patients examined in an earlier disease-stage.

ECV was mildly increased both in the current study and in previous WD studies (26–28). Elevated ECV indicates diffuse fibrosis or infiltration (25) and since copper accumulates intracellularly (33), the increased ECV should reflect diffuse fibrosis. This theory is supported by autopsies showing cardiac hypertrophy, interstitial and replacement fibrosis, intramyocardial small vessel sclerosis and inflammation in deceased newly diagnosed or chronic WD patients, some with sudden death (34). Interestingly, in hypertrophic cardiomyopathy (HCM) similarly characterized by hypertrophy, interstitial and replacement fibrosis and dysplastic intramyocardial arterioles, decoppering improves LV GLS and reduces T1, ECV and LVM by reducing extracellular matrix (ECM), meaning reducing diffuse fibrosis (35). Therefore, as the stable WD patients in our study had only a slight increase in ECV and no differences were found in native T1, LVM or LV GLS, this may indicate successful decoppering.

### Fibrosis, hypertrophy and impaired stress perfusion

We showed a high prevalence of RVIP-LGE, and three patients had basal lateral or inferolateral non-ischemic LGE, of which two had elevated native T2. Similarly, focal fibrosis has previously been shown in WD, most often in the RVIP or interventricular septum (27–29, 36), together with increased LVM without LV hypertrophy (LVH) and with impaired RV function (29, 36). In contrast, LVM and RV function were unaffected in the current study. In HCM, RVIP-LGE is the most common LGE-pattern and associated with lower LVEF (37). Similarly, our WD patients had lower LVEF. RVIP-LGE represents intermediate HCM and has been shown with impaired LV GLS measured by Echo, and reduced stress perfusion and MPR measured by positron emission tomography (38). Additionally, reduced stress perfusion and MPR have been shown in HCM using stress perfusion CMR, with lower stress perfusion in more hypertrophied and fibrotic myocardium (39, 40). Furthermore, in the sphingolipid-storage disease Fabry which causes inflammation, fibrosis and LVH, reduced stress perfusion has been shown even before LVH, indicating that reduced stress perfusion is an early disease marker in this hypertrophic cardiomyopathy (41). Moreover, impaired stress perfusion and LV GLS have been shown pre-LVH in Fabry (42). While LV GLS was unaffected, we also demonstrated reduced stress perfusion and MPR in WD patients. Inability to increase myocardial perfusion during stress indicate presence of CMD in the absence of epicardial coronary disease, which in this setting may be caused by oxidative stress, inflammation, fibrosis, small vessel sclerosis or dysplasia with capillary rarefaction and hypertrophy (43). Therefore, the increased ECV and reduced stress perfusion and MPR may reflect early cardiac involvement with diffuse fibrosis leading to CMD and mild cardiac dysfunction developing prior to more pronounced LVH, which we theorize to be secondary to inflammation caused by copper exposure.

### Cardiac dysfunction, fibrosis and impaired stress perfusion

LVEF was within normal range but slightly lower, and LV GCS was slightly increased in our WD patients. LV GRS and LV GLS were unaffected, and LV GCS and LV GRS correlated with stress perfusion in WD patients. Previous studies have shown no differences in strain in WD (26, 27), but also impaired LV GRS (36) and similar to our results, impaired LV GCS (28).

Impaired RV function has been reported (29, 36), which was not present in the current study. In HCM with preserved LVEF, impaired LV GLS and LV GCS have been reported with LV GCS contributing twice as much to LVEF as LV GLS owing to compensation by midwall circumferential fibers compared to subendocardial longitudinal fibers that are more sensitive to cardiac disease (44). LV GCS becomes impaired later than LV GLS and impaired LV GCS therefore indicates more severe transmural myocardial injury (45). Thus, it is surprising that LV GCS but not LV GLS or LV GRS were impaired. However, segmental circumferential strain may identify and predict fibrotic segments in HCM (46, 47). Perhaps minor GCS impairment reflects early fibrosis with a particular pattern, occurring before major hypertrophy and thus impairment in cardiac function, too minor or diffuse to cause focal LGE. Moreover, impaired LV strain have been shown in HCM, with impaired circumferential and radial strain in segments with perfusion defects and LGE (48). Additionally, HCM patients with hypertrophy have more perfusion defects, and segments with perfusion defects have higher T1, T2, ECV, LGE and impaired circumferential and radial strain (49). In patients with heart failure with preserved ejection fraction, increased ECV has been shown to correlate with impaired LV GLS, LV GCS and LV GRS (50). This further supports our theory, that the WD patients may have mild diffuse fibrosis which may lead to subclinical impairment in perfusion and myocardial function.

### Limitations

The current study is limited by its single-center design, small sample size and limited clinical data. Biopsy of liver and myocardium was not performed, therefore the CMR findings cannot be correlated to histopathology. The patients were relatively young and without cardiac symptoms, therefore this cohort likely reflect early myocardial involvement with discrete findings on CMR. Further studies in greater patient populations are needed to reproduce the results and further our understanding of myocardial involvement in WD.

## Conclusions

In this small mechanistic study, WD patients on stable treatment without cardiac symptoms have early signs of diffuse fibrosis, CMD and systolic dysfunction, shedding light on pathophysiological mechanisms of cardiac dysfunction in copper accumulation.

## Clinical Perspectives

### Competency in medical knowledge

Even relatively young WD patients without cardiac symptoms may have signs of cardiac involvement including diffuse fibrosis, CMD and mild systolic dysfunction.

### Translational outlook

Future CMR studies in WD patients should explore cardiac manifestations in larger cohorts in multi-center settings over a broader clinical spectrum, together with clinical data and outcome data.

## Data Availability

Upon reasonable request, data supporting the findings in this study are available from the corresponding author.

## Abbreviations

CMR: Cardiovascular Magnetic Resonance Imaging
ECV: Extracellular Volume
LGE: Late Gadolinium Enhancement
LV GCS: Left Ventricular Global Circumferential Strain
LV GLS: Left Ventricular Global Longitudinal Strain
LV GRS: Left Ventricular Global Radial Strain
MPR: Myocardial Perfusion Reserve
RVIP: Right Ventricular Insertion Point
SAQ-7: Seattle Angina Questionnaire-7
WD: Wilson Disease

## Declarations

### Ethical approval and consent to participate

The Swedish Ethical Review Authority has granted ethical approval for the study (DNR 2019- 06287, with amendments 2019-06287, 2021-05508-02, 2022-06995-02 and for volunteers 2015/2106–31/1 and 2014/131-31/1, 2017/2415-32/1, 2021-06837-02). All participants provided written informed consent.

### Consent for publication

All participants provided written informed consent with regards to publication of individual data on group level and to publish anonymized images. Consent forms are documented in the clinical notes of the participants and original forms are held in our research office and are available for the Editor-in-Chief for review.

### Authors’ contributions

Steffen Johansson recruited participants, participated in image acquisition, performed image, data and statistical analysis as well as drafted the manuscript. Fogarasi participated in image, data and statistical analysis. Nickander and Kindmark designed the study. Nickander participated in image acquisition, and supervised image, data and statistical analysis. All authors read, revised and approved the final manuscript.

---

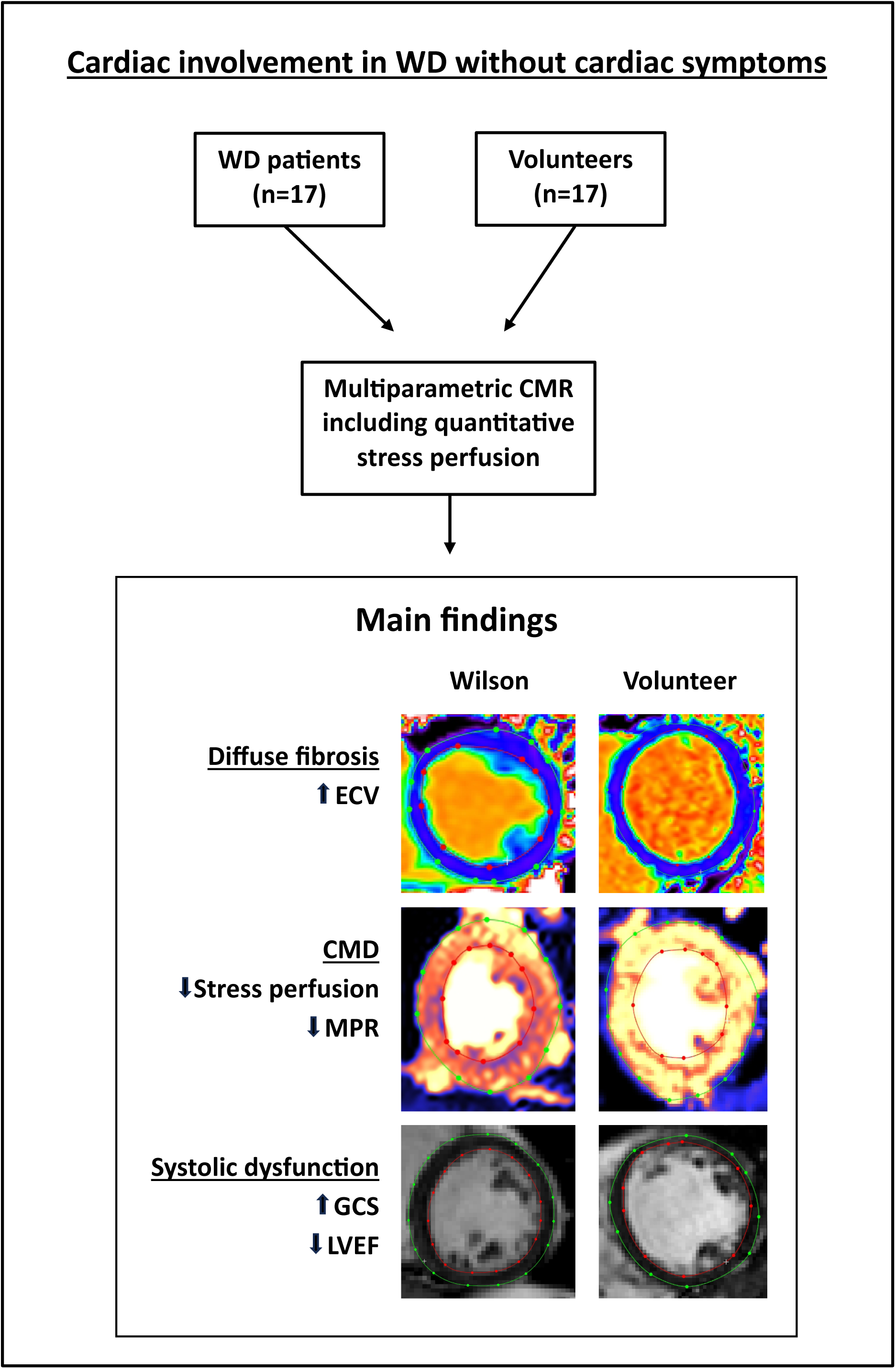
Central illustration. The main findings of our study comparing WD patients without cardiac symptoms with healthy volunteers using multiparametric CMR including stress perfusion are higher global ECV, reduced stress perfusion and MPR as well as impaired GCS and lower LVEF in WD patients. Abbreviations: WD = Wilson disease; CMD = coronary microvascular dysfunction; CMR = cardiovascular magnetic resonance imaging; ECV = extracellular volume; MPR = myocardial perfusion reserve; GCS = global circumferential strain; LVEF = left ventricular ejection fraction.

## References

1. Mulligan C, Bronstein JM. Wilson Disease: An Overview and Approach to Management. Neurol Clin. 2020;38(2):417–32.

2. Kuan P. Cardiac Wilson’s disease. Chest. 1987;91(4):579–83.

3. Grandis DJ, Nah G, Whitman IR, Vittinghoff E, Dewland TA, Olgin JE, et al. Wilson’s Disease and Cardiac Myopathy. Am J Cardiol. 2017;120(11):2056–60.

4. Buksińska-Lisik M, Litwin T, Pasierski T, Członkowska A. Cardiac assessment in Wilson’s disease patients based on electrocardiography and echocardiography examination. Arch Med Sci. 2019;15(4):857–64.

5. Karhan AN, Aykan HH, Gümüş E, Dönmez Y, Alehan D, Özkutlu S, et al. Assessment of cardiac function and electrocardiographic findings in patients with Wilson’s disease. Cardiol Young. 2019;29(9):1183–8.

6. Hlubocká Z, Marecek Z, Linhart A, Kejková E, Pospísilová L, Martásek P, et al. Cardiac involvement in Wilson disease. J Inherit Metab Dis. 2002;25(4):269–77.

7. Bobbio E, Forsgard N, Oldfors A, Szamlewski P, Bollano E, Andersson B, et al. Cardiac arrest in Wilson’s disease after curative liver transplantation: a life-threatening complication of myocardial copper excess? ESC Heart Fail. 6. England: © 2019 The Authors. ESC Heart Failure published by John Wiley & Sons Ltd on behalf of the European Society of Cardiology.; 2019. p. 228-31.

8. Chevalier K, Benyounes N, Obadia MA, Van Der Vynckt C, Morvan E, Tibi T, et al. Cardiac involvement in Wilson disease: Review of the literature and description of three cases of sudden death. J Inherit Metab Dis. 2021;44(5):1099–112.

9. Zhou W, Sin J, Yan AT, Wang H, Lu J, Li Y, et al. Qualitative and Quantitative Stress Perfusion Cardiac Magnetic Resonance in Clinical Practice: A Comprehensive Review. Diagnostics (Basel). 2023;13(3).

10. European Association for Study of L. EASL Clinical Practice Guidelines: Wilson’s disease. J Hepatol. 2012;56(3):671-85.

11. Nickander J, Themudo R, Thalén S, Sigfridsson A, Xue H, Kellman P, et al. The relative contributions of myocardial perfusion, blood volume and extracellular volume to native T1 and native T2 at rest and during adenosine stress in normal physiology. J Cardiovasc Magn Reson. 2019;21(1):73.

12. Steffen Johansson R, Tornvall P, Sörensson P, Nickander J. Reduced stress perfusion in myocardial infarction with nonobstructive coronary arteries. Sci Rep. 2023;13(1):22094.

13. Chan PS, Jones PG, Arnold SA, Spertus JA. Development and validation of a short version of the Seattle angina questionnaire. Circ Cardiovasc Qual Outcomes. 2014;7(5):640–7.

14. Bassingthwaighte JB, Wang CY, Chan IS. Blood-tissue exchange via transport and transformation by capillary endothelial cells. Circ Res. 1989;65(4):997–1020.

15. Kellman P, Hansen MS, Nielles-Vallespin S, Nickander J, Themudo R, Ugander M, et al. Myocardial perfusion cardiovascular magnetic resonance: optimized dual sequence and reconstruction for quantification. J Cardiovasc Magn Reson. 2017;19(1):43.

16. Kellman P, Wilson JR, Xue H, Ugander M, Arai AE. Extracellular volume fraction mapping in the myocardium, part 1: evaluation of an automated method. J Cardiovasc Magn Reson. 2012;14(1):63.

17. Arheden H, Saeed M, Higgins CB, Gao DW, Bremerich J, Wyttenbach R, et al. Measurement of the distribution volume of gadopentetate dimeglumine at echo-planar MR imaging to quantify myocardial infarction: comparison with 99mTc-DTPA autoradiography in rats. Radiology. 1999;211(3):698–708.

18. Heiberg E, Sjögren J, Ugander M, Carlsson M, Engblom H, Arheden H. Design and validation of Segment--freely available software for cardiovascular image analysis. BMC Med Imaging. 2010;10:1.

19. Queiros S, Morais P, Barbosa D, Fonseca JC, Vilaca JL, D’Hooge J. MITT: Medical Image Tracking Toolbox. IEEE Trans Med Imaging. 2018;37(11):2547–57.

20. Morais P, Marchi A, Bogaert JA, Dresselaers T, Heyde B, D’Hooge J, et al. Cardiovascular magnetic resonance myocardial feature tracking using a non-rigid, elastic image registration algorithm: assessment of variability in a real-life clinical setting. J Cardiovasc Magn Reson. 2017;19(1):24.

21. Hundley WG, Bluemke D, Bogaert JG, Friedrich MG, Higgins CB, Lawson MA, et al. Society for Cardiovascular Magnetic Resonance guidelines for reporting cardiovascular magnetic resonance examinations. J Cardiovasc Magn Reson. 2009;11:5.

22. Mosteller RD. Simplified calculation of body-surface area. N Engl J Med. 1987;317(17):1098.

23. Nickander J, Themudo R, Sigfridsson A, Xue H, Kellman P, Ugander M. Females have higher myocardial perfusion, blood volume and extracellular volume compared to males - an adenosine stress cardiovascular magnetic resonance study. Sci Rep. 2020;10(1):10380.

24. Kanda T, Nakai Y, Aoki S, Oba H, Toyoda K, Kitajima K, et al. Contribution of metals to brain MR signal intensity: review articles. Jpn J Radiol. 2016;34(4):258–66.

25. Messroghli DR, Moon JC, Ferreira VM, Grosse-Wortmann L, He T, Kellman P, et al. Correction to: Clinical recommendations for cardiovascular magnetic resonance mapping of T1, T2, T2* and extracellular volume: A consensus statement by the Society for Cardiovascular Magnetic Resonance (SCMR) endorsed by the European Association for Cardiovascular Imaging (EACVI). J Cardiovasc Magn Reson. 2018;20(1):9.

26. Deng W, Zhang J, Zhao R, Qian Y, Han Y, Yu Y, et al. T1 Mapping Values May Be Associated with Early Myocardial Involvement in Young Patients with Wilson Disease. Radiol Cardiothorac Imaging. 2022;4(6):e220145.

27. Deng W, Zhang J, Jia Z, Pan Z, Wang Z, Xu H, et al. Myocardial involvement characteristics by cardiac MR imaging in neurological and non-neurological Wilson disease patients. Insights Imaging. 2024;15(1):24.

28. Salatzki J, Mohr I, Heins J, Cerci MH, Ochs A, Paul O, et al. The impact of Wilson disease on myocardial tissue and function: a cardiovascular magnetic resonance study. J Cardiovasc Magn Reson. 2021;23(1):84.

29. Quick S, Weidauer M, Heidrich FM, Sveric K, Reichmann H, Ibrahim K, et al. Cardiac Manifestation of Wilson’s Disease. J Am Coll Cardiol. 2018;72(22):2808–9.

30. Dusek P, Litwin T, Członkowska A. Neurologic impairment in Wilson disease. Ann Transl Med. 2019;7(Suppl 2):S64.

31. Meloni A, Carnevale A, Gaio P, Positano V, Passantino C, Pepe A, et al. Liver T1 and T2 mapping in a large cohort of healthy subjects: normal ranges and correlation with age and sex. MAGMA. 2024;37(1):93–100.

32. Isaak A, Praktiknjo M, Jansen C, Faron A, Sprinkart AM, Pieper CC, et al. Myocardial Fibrosis and Inflammation in Liver Cirrhosis: MRI Study of the Liver-Heart Axis. Radiology. 2020;297(1):51–61.

33. Ralle M, Huster D, Vogt S, Schirrmeister W, Burkhead JL, Capps TR, et al. Wilson disease at a single cell level: intracellular copper trafficking activates compartment-specific responses in hepatocytes. J Biol Chem. 2010;285(40):30875–83.

34. Factor SM, Cho S, Sternlieb I, Scheinberg IH, Goldfischer S. The cardiomyopathy of Wilson’s disease. Myocardial alterations in nine cases. Virchows Arch A Pathol Anat Histol. 1982;397(3):301–11.

35. Reid A, Miller C, Farrant JP, Polturi R, Clark D, Ray S, et al. Copper chelation in patients with hypertrophic cardiomyopathy. Open Heart. 2022;9(1).

36. Zhang K, Reuner U, Hempel C, Speiser U, Ibrahim K, Heinzel FR, et al. Evaluation of Myocardial Strain Using Cardiac Magnetic Resonance in Patients with Wilson’s Disease. J Clin Med. 2021;10(2).

37. Zhu Y, Park EA, Lee W, Kim HK, Chu A, Chung JW, et al. Extent of late gadolinium enhancement at right ventricular insertion points in patients with hypertrophic cardiomyopathy: relation with diastolic dysfunction. Eur Radiol. 2015;25(4):1190–200.

38. Bravo PE, Luo HC, Pozios I, Zimmerman SL, Corona-Villalobos CP, Sorensen L, et al. Late gadolinium enhancement confined to the right ventricular insertion points in hypertrophic cardiomyopathy: an intermediate stage phenotype? Eur Heart J Cardiovasc Imaging. 2016;17(3):293–300.

39. Petersen SE, Jerosch-Herold M, Hudsmith LE, Robson MD, Francis JM, Doll HA, et al. Evidence for microvascular dysfunction in hypertrophic cardiomyopathy: new insights from multiparametric magnetic resonance imaging. Circulation. 2007;115(18):2418–25.

40. Camaioni C, Knott KD, Augusto JB, Seraphim A, Rosmini S, Ricci F, et al. Inline perfusion mapping provides insights into the disease mechanism in hypertrophic cardiomyopathy. Heart. 2020;106(11):824–9.

41. Knott KD, Augusto JB, Nordin S, Kozor R, Camaioni C, Xue H, et al. Quantitative Myocardial Perfusion in Fabry Disease. Circ Cardiovasc Imaging. 2019;12(7):e008872.

42. Augusto JB, Johner N, Shah D, Nordin S, Knott KD, Rosmini S, et al. The myocardial phenotype of Fabry disease pre-hypertrophy and pre-detectable storage. Eur Heart J Cardiovasc Imaging. 2021;22(7):790–9.

43. Vancheri F, Longo G, Vancheri S, Henein M. Coronary Microvascular Dysfunction. J Clin Med. 2020;9(9).

44. Stokke TM, Hasselberg NE, Smedsrud MK, Sarvari SI, Haugaa KH, Smiseth OA, et al. Geometry as a Confounder When Assessing Ventricular Systolic Function: Comparison Between Ejection Fraction and Strain. J Am Coll Cardiol. 2017;70(8):942–54.

45. Bansal M, Sengupta PP. Longitudinal and circumferential strain in patients with regional LV dysfunction. Curr Cardiol Rep. 2013;15(3):339.

46. Sharifian M, Rezaeian N, Asadian S, Mohammadzadeh A, Nahardani A, Kasani K, et al. Efficacy of Novel Noncontrast Cardiac Magnetic Resonance Methods in Indicating Fibrosis in Hypertrophic Cardiomyopathy. Cardiol Res Pract. 2021;2021:9931136.

47. She J, Zhao S, Chen Y, Zeng M, Jin H. Detecting Regional Fibrosis in Hypertrophic Cardiomyopathy: The Utility of Myocardial Strain Based on Cardiac Magnetic Resonance. Acad Radiol. 2023;30(2):230–8.

48. Garcia Brás P, Rosa SA, Cardoso I, Branco LM, Galrinho A, Gonçalves AV, et al. Microvascular Dysfunction Is Associated With Impaired Myocardial Work in Obstructive and Nonobstructive Hypertrophic Cardiomyopathy: A Multimodality Study. J Am Heart Assoc. 2023;12(8):e028857.

49. Garcia Brás P, Aguiar Rosa S, Thomas B, Fiarresga A, Cardoso I, Pereira R, et al. Associations between perfusion defects, tissue changes and myocardial deformation in hypertrophic cardiomyopathy, uncovered by a cardiac magnetic resonance segmental analysis. Rev Port Cardiol. 2022;41(7):559–68.

50. Zhang Y, Zhang X, Wang Y, Hu X, Wang B, Yang J, et al. Relationship between diffuse fibrosis assessed by CMR and depressed myocardial strain in different stages of heart failure. Eur J Radiol. 2023;164:110848.

