## Appendix for "Diffuse fibrosis, coronary microvascular dysfunction and systolic dysfunction in Wilson disease"

**Table 1. Intra- and inter-observer agreement**

| | **Intra-observer agreement** | **Inter-observer agreement** |
|---|---|---|
| **Native T1** | 0.99, $p$<0.001 | 0.99, $p$<0.001 |
| **Native T2** | 0.98, $p$<0.001 | 0.98, $p$<0.001 |
| **ECV** | 1.00, $p$<0.001 | 0.91, $p$<0.001 |
| **Perfusion rest** | 1.00, $p$<0.001 | 1.00, $p$<0.001 |
| **Perfusion stress** | 1.00, $p$<0.001 | 0.99, $p$<0.001 |
| **MPR** | 0.99, $p$<0.001 | 0.99, $p$<0.001 |
| **LVEF** | 0.89, $p$=0.002 | 0.85, $p$<0.001 |
| **LV GLS** | 0.95, $p$<0.001 | 0.94, $p$<0.001 |
| **LV GCS** | 0.84, $p$=0.008 | 0.92, $p$<0.001 |
| **LV GRS** | 0.92, $p$<0.001 | 0.82, $p$<0.001 |

Intra- and inter-observer agreement presented as intraclass correlation coefficient (ICC) together with p-values. Abbreviations: ECV = extracellular volume; MPR = myocardial perfusion reserve; LVEF = left ventricular ejection fraction; LV GLS = left ventricular global longitudinal strain; LV GCS = left ventricular global circumferential strain; LV GRS = left ventricular global radial strain.

**Table 2. RV volumes and function**

| CMR findings | WD patients (n=17) | Volunteers (n=17) | *p* |
|---|---|---|---|
| **RVEDV (ml)** | 170±41 | 165±35 | 0.68 |
| **RVEDVi(ml/m$^2$)** | 88±19 | 89±15 | 0.93 |
| **RVESV (ml)** | 64±21 | 58±21 | 0.43 |
| **RVESVi (ml/m$^2$)** | 33±11 | 31±11 | 0.63 |
| **RVSV (ml)** | 107±29 | 107±26 | 0.97 |
| **RVSVi (ml/m$^2$)** | 55±12 | 57±10 | 0.55 |
| **RVEF (%)** | 63±9 | 65±9 | 0.40 |
| **RV GLS (%)** | -18±4 | -20±3 | 0.16 |
| **RV AVPD (mm)** | -19±4 | -20±3 | 0.63† |

CMR findings are presented as mean ± standard deviation. P-values denotes the independent t-test, p-values marked † denotes the Mann Whitney U test. Abbreviations: CMR = cardiovascular magnetic resonance imaging; RV = right ventricle; RVEDV = right ventricular end-diastolic volume; RVESV = right ventricular end-systolic volume; RVSV = right ventricular stroke volume; RVEF = right ventricular ejection fraction; RV GLS = right ventricular global longitudinal strain; RV AVPD = right ventricle atrioventricular plane displacement; WD = Wilson disease.
